# Prevalence of bias attributable to composite outcome in clinical trials: a systematic review

**DOI:** 10.1101/2024.07.18.24310633

**Authors:** José Mário Nunes da Silva, Juliana Ferreira Souza Conceição, Paula C. Ramírez, Christian Leonardo Diaz-León, Fredi Alexander Diaz-Quijano

**Author notes:** Corresponding author: José Mário Nunes da Silva, Graduate Program in Epidemiology, School of Public Health, University of São Paulo, Av. Dr. Arnaldo, 715, Cerqueira César, São Paulo, SP, 01246-904, Brazil.

## Abstract

**Objective:** To investigate the prevalence of bias attributable to composite outcome (BACO) in clinical trials.

**Study design and setting:** We searched PubMed for randomized clinical trials where the primary outcome was a binary composite that included all-cause mortality among its components from January 1, 2019, to December 31, 2020. For each trial, the BACO index was calculated to assess the correspondence between effects on the composite outcome and that on mortality. This systematic review was registered in PROSPERO (CRD42021229554).

**Results:** After screening 1,076 citations and 171 full-text articles, 91 studies were included from 13 different medical areas. The prevalence of significant or suggestive BACO among the 91 included articles was 25.2% (n=23), including 12 with p<0.005 and 11 with p between 0.005 and <0.05. We observed that in 17 (73.9%) of these 23 studies, the BACO index value was between zero and <1, indicating an underestimation of the effect. The other six studies showed negative values (26.1%), indicating an inversion of the association with mortality. None of the studies showed significant overestimation of the association attributable to the composite outcome.

**Conclusion:** These findings highlight the need to predefine guidelines for interpreting effects on composite endpoints based on objective criteria such as the BACO index.

**What is new?:** *Key Findings:*

- The study found that 25.2% of the included clinical trials exhibited significant or suggestive bias attributable to composite outcomes (BACO).
- In 73.9% of these cases, the BACO index was less than 1, indicating an underestimation of the effect. 26.1% of the studies showed an inversion of the association with mortality.
- No significant overestimation of the association due to composite outcomes was observed.

*What This Adds to What Was Known?:*

- This study contributes to the existing knowledge by quantifying the prevalence of bias attributable to composite outcomes in clinical trials.
- It highlights that a significant proportion of trials may underestimate the effect or even show an inversion of the association with mortality when composite outcomes are used.
- This finding emphasizes the need for careful consideration and objective criteria, like the BACO index, in the design and interpretation of clinical trials involving composite outcomes.

*What Is the Implication and What Should Change Now?:*

- Researchers and clinicians should be cautious about relying solely on composite outcomes without assessing the potential biases they introduce.
- The study suggests a need for predefined guidelines and objective criteria, such as the BACO index, for interpreting the effects of composite outcomes.

## 1. Introduction

Composite outcomes are often used in randomized trials to assess the efficacy of a new intervention compared to standard treatment [1,2]. Their use involves analyzing a greater number of outcomes over shorter follow-up periods. Typically, this is expected to increase the power of the study, reduce costs, and provide a quicker response to a research question [3]. However, composite outcomes can lead to misleading conclusions when the individual components, which may vary in importance and frequency, are affected differently or oppositely by the interventions being evaluated [4,5].

In this context, the Bias Attributable to Composite Outcome (BACO) Index is a recently developed strategy that aids in interpreting the effects on a composite outcome [6]. This index corresponds to the ratio of the logarithms of the measures of association between the composite outcome and mortality. BACO index values different from one indicate that using a composite outcome affects the prognosis as follows: overestimated (BACO index > 1), underestimated (BACO index between 0 and < 1), or inverted (BACO index < 0), using the effect on mortality as the reference point [6].

Despite the frequent use of composite outcomes, especially in cardiovascular clinical trials, the frequency and direction of BACO have not been widely quantified. Therefore, we aim to investigate the prevalence of BACO in clinical trials published in PubMed between 2019 and 2020.

## 2. Methods

### 2.1 Protocol and registration

We conducted the review according to the guidelines of the Preferred Reporting Items for Systematic Reviews and Meta-Analyses (PRISMA) [7]. We registered the protocol in PROSPERO (registration number: CRD42021229554).

### 2.2 Eligibility Criteria

#### 2.2.1 Inclusion criteria

We included randomized clinical trials where the primary outcome was a binary composite outcome that included all-cause mortality among its components [6].

#### 2.2.2 Exclusion criteria

We excluded cluster randomized trials, secondary analyses, subgroup analyses, and studies with fewer than five fatal events. Additionally, we did not include four articles that lacked data on the frequency of composite outcomes or mortality.

### 2.3 Search strategy

We searched PubMed for articles published electronically in English, Portuguese, and Spanish between January 2019 and December 2020 (updated on April 5, 2021). We used the following terms: Composite AND primary AND (endpoint OR outcome OR (“end-point”)) AND (mortality OR death) AND (randomized OR randomised) AND (trial).

### 2.4 Selection of studies

Pairs of two independent reviewers (J.F.S.C and P.C.R; J.M.N.S and C.L.D.L) screened titles and abstracts of all citations retrieved during the literature search. Subsequently, reviewers read potentially eligible articles in full to determine if they met the eligibility criteria. Any discrepancies between reviewers that arose at each stage of the study selection process were resolved through consensus and, if needed, arbitration by a third reviewer (F.A.D.Q). Prior to both steps of study selection, we conducted a pilot-tested using random samples of 10 full articles.

### 2.5 Data extraction and management

We used a pre-defined standardized protocol where two independent reviewers (J.F.S.C and P.C.R) extracted data from included studies, compared information, and resolved disagreements through discussion. We extracted the following data: article title, year of publication, first author’s name, journal published, randomization and blinding process, follow-up period, experimental and control group interventions, sample size in each group, sample loss, components of the composite outcome, number of composite endpoints and deaths, measure of association used, and information on intention-to-treat analysis. We also recorded whether the study’s conclusion was based on the composite outcome and if authors addressed discrepancies between the composite outcome result and mortality, as well as protocol registration on a platform.

### 2.6 Data analysis

We calculated the BACO index (Bias Attributable to Composite Outcome) (6) defined as [6]: where <p_C_ e <p_d_ are the relative risks for the composite outcome (RRc) and mortality (RRd), respectively.

A BACO index equal to one indicates no bias attributable to using a composite outcome, with mortality as the reference measure of association [6].

We set a predefined significance level of 0.005. However, we used the term suggestive for p-values between 0.005 and 0.05. The 95% confidence intervals for the BACO index and hypothesis testing were conducted following the methods described in the original study [6]. We performed the analyses using Microsoft Office Excel 2019 (Microsoft Corporation, Redmond, Washington).

### 2.7 Risk of bias in individual studies

For studies suggestive of BACO or with significant BACO, we conducted a bias assessment by two independent reviewers (J.F.S.C and P.C.R) using the updated version of the Cochrane tool (RoB2) [8]. This tool assesses bias risk across the following domains: bias arising from the randomization process, bias due to deviations from intended interventions, bias from missing outcome data, outcome measurement bias, and bias in the reporting of results. Discrepancies were reassessed by a third reviewer (J.M.N.S), and consensus was reached through discussion.

## 3. Results

### 3.1 Search results

The initial search provided us a total of 1,076 studies. After reviewing titles and abstracts, we excluded 905 studies. Of the 171 eligible full-text articles, 123 met the study’s inclusion criteria. However, we excluded 32 studies due to fewer than 5 total deaths. Ultimately, we selected 91 studies for this review (Fig. 1).

**Figure 1.**
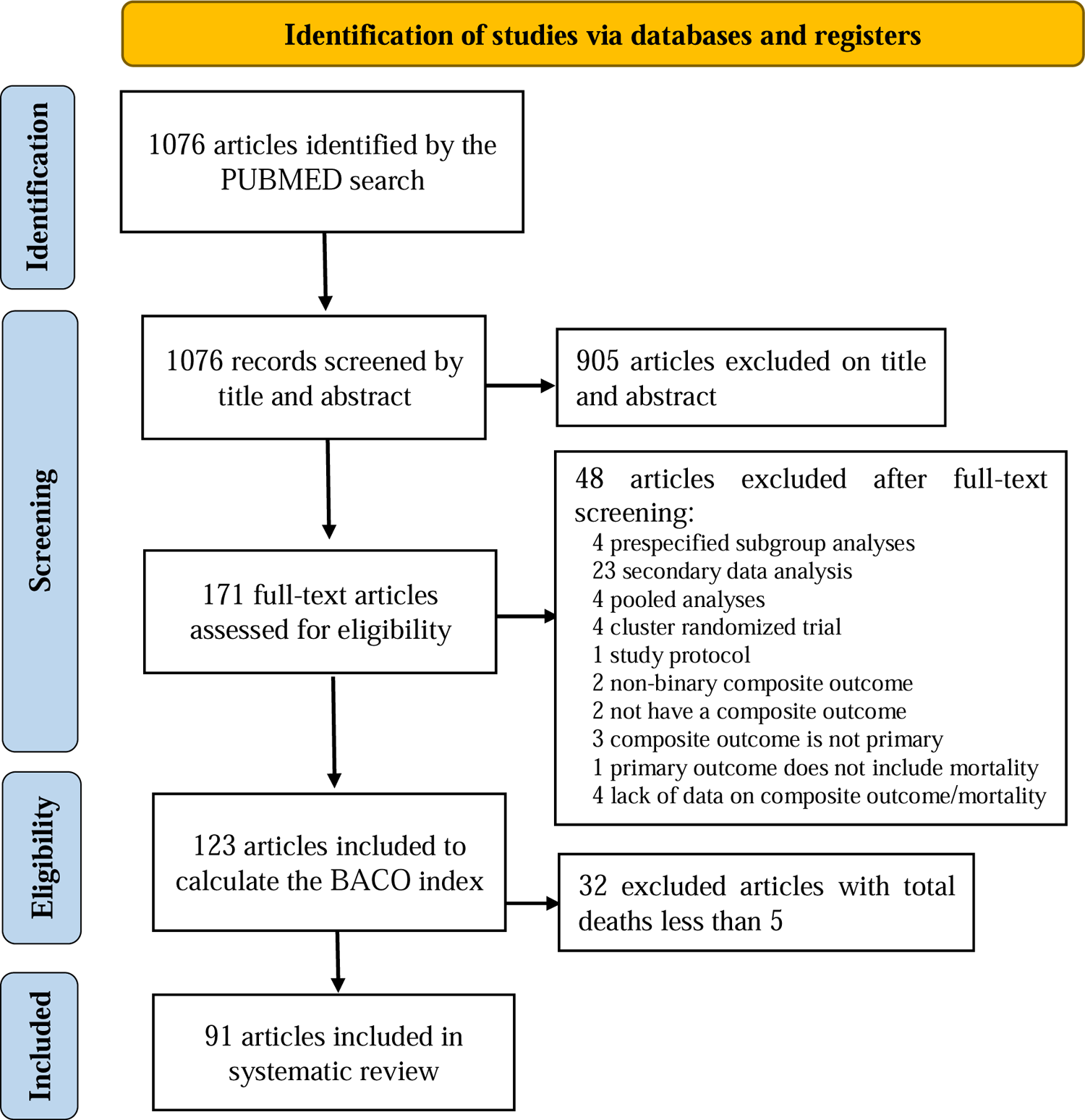
Flow chart of the study identification and selection.

### 3.2 Study characteristics

All articles were written in English and were published in 44 different journals covering 13 different thematic areas, according to Scopus classification. The most prevalent categories were Cardiology and Cardiovascular Medicine (37/91; 40.7%) and Medicine (various, 33/91; 36.2%). The manuscripts were most frequently published in The New England Journal of Medicine (17/91; 18.7%), Circulation (12/91; 13.2%), and JAMA (10/91; 11%) (Table 1).

**Table 1.** Characteristics of the articles included (n=91).

| **Study characteristic** | **N** | **%** |
| --- | --- | --- |
| Scopus subject category |  |  |
| Cardiology and Cardiovascular Medicine | 37 | 40.7 |
| Medicine (miscellaneous) | 33 | 36.2 |
| General Medicine | 5 | 5.5 |
| Pediatrics, Perinatology and Child Health | 4 | 4.4 |
| Nephrology | 2 | 2.2 |
| Critical Care and Intensive Care Medicine | 2 | 2.2 |
| Pulmonary and Respiratory Medicine | 2 | 2.2 |
| Others | 6 | 6.6 |
| Publication Journal |  |  |
| The New England Journal of Medicine | 17 | 18.7 |
| Circulation | 12 | 13.2 |
| JAMA | 10 | 11 |
| The Lancet | 5 | 5.5 |
| American Heart Journal | 3 | 3.3 |
| European Heart Journal | 3 | 3.3 |
| Journal of the American College of Cardiology | 3 | 3.3 |
| Others | 38 | 41.7 |
| Intention to treat analysis |  |  |
| Yes | 84 | 92.3 |
| Blinding |  |  |
| Yes | 76 | 83.5 |
| Platform protocol registration | 84 | 92.3 |
| Yes |  |  |
| Random sequence Generation |  |  |
| Specific software | 64 | 70.3 |
| Permuted block randomization | 12 | 13.2 |
| Random number table | 2 | 2.2 |
| Raffle | 1 | 1.1 |
| Unspecified | 12 | 13.2 |

The majority (84/91; 92.3%) of included articles had registered protocols, and the same number conducted intention-to-treat analysis. Masked intervention was evaluated in most studies (76/91; 83.5%), and allocation was based on specific software randomization sequences in 64/91 studies (70.3%). Six articles reported discrepancies between the composite outcome association and mortality (6/91; 6.6%), and conclusions were not based on the composite outcome in two manuscripts (2/91; 2.2%).

### 3.3 Prevalence of BACO

Out of the 91 included articles, 23 (25.2%) had a significant or suggestive BACO, including 12 with p<0.005 and 11 with p between 0.005 and <0.05 (Tables 2 and 3). These 23 studies included a total of 21,285 participants in the experimental group, with sample sizes ranging from 46 to 5,523 patients, a median of 417 (IQR: 182-1,050); presenting 24 to 975 composite outcomes, with a median of 89 (IQR: 69-203); and 2 to 119 deaths, with a median of 15 (IQR: 7-36). In the control groups, there were a total of 20,048 participants, with sample sizes ranging from 54 to 5,493, a median of 409 (IQR: 190-1,038). In these groups, the number of composite events ranged from 19 to 924, with a median of 88 (IQR: 60-176); and fatal events ranged from 1 to 100, with a median of 11 (IQR: 6-38) (Table 2).

**Table 2.**
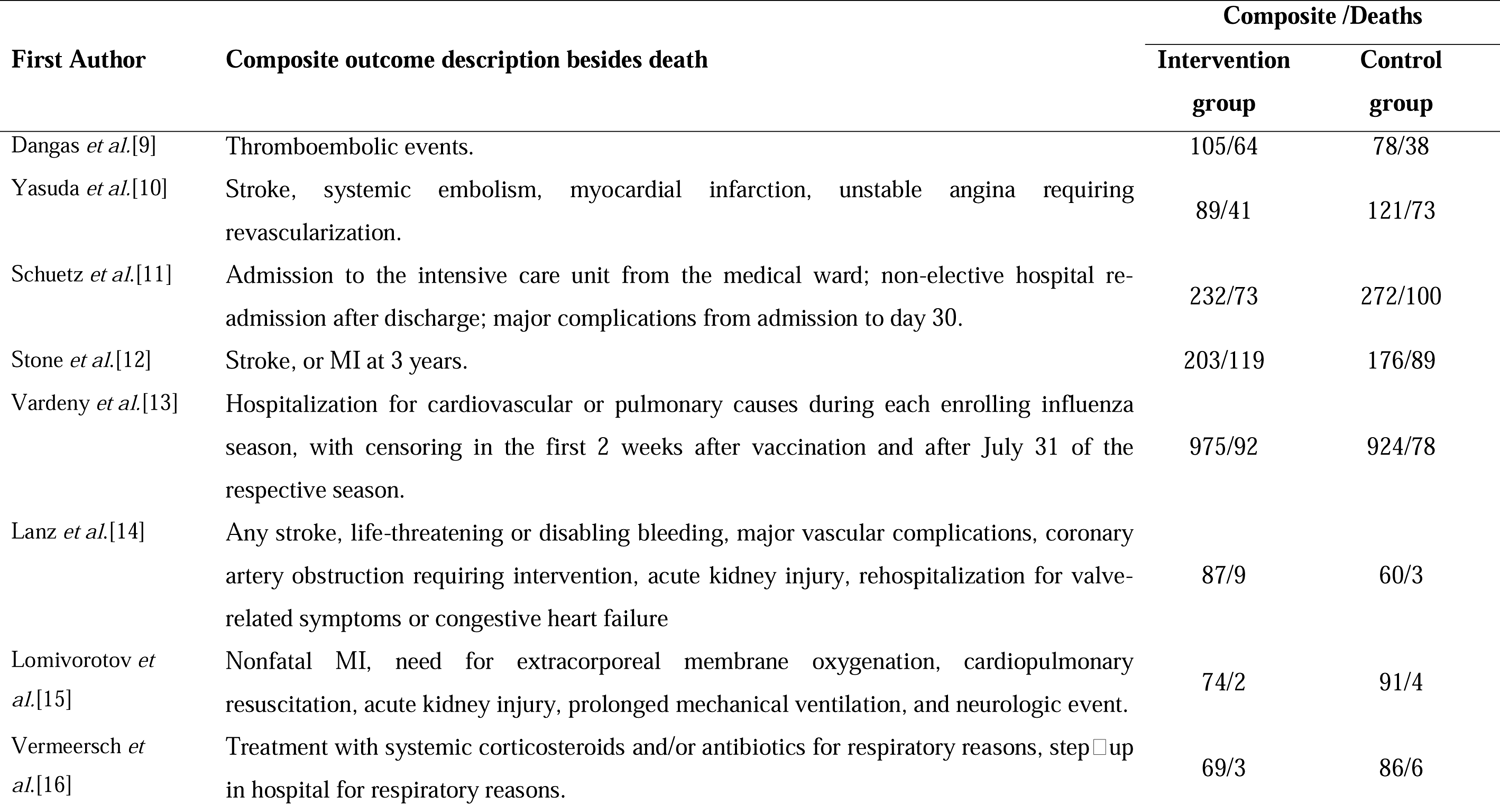
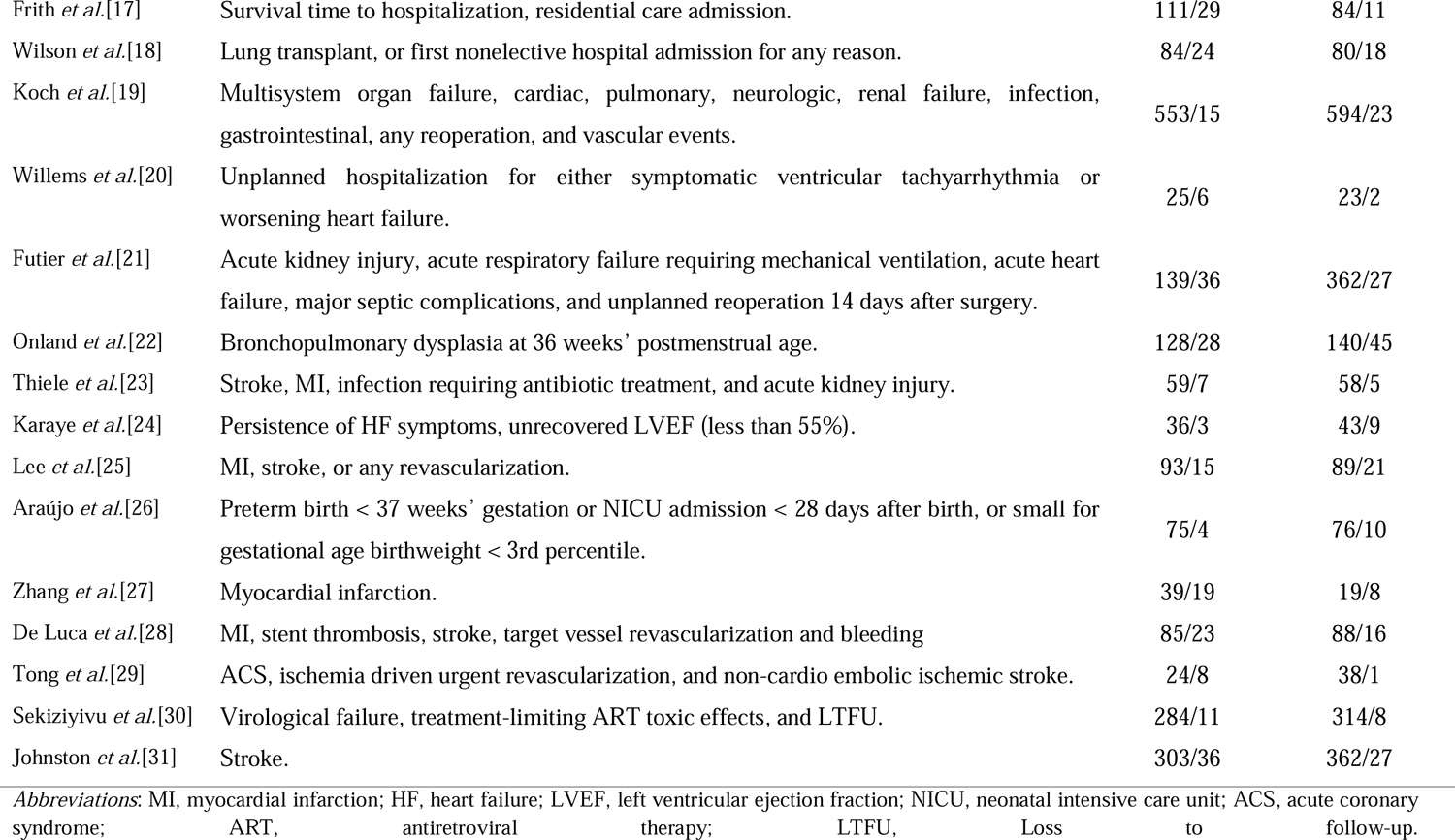
General description of the study population and outcomes of clinical trials selected (n=23).

**Table 3.** Relative risks of composite and death and BACO index in clinical trials (n=23).

| First author | RR<sub>c</sub> (95% CI) | RR<sub>d</sub> (95% CI) | BACO index<br>(95% CI) | p value<sup>a</sup> |
| --- | --- | --- | --- | --- |
| Dangas *et al.*[9] | 1.33 (1.01 to 1.76) | 1.67 (1.13 to 2.46) | 0.56 (0.18 to 0.95) | 0.025 |
| Yasuda *et al.*[10] | 0.74 (0.57 to 0.95) | 0.56 (0.39 to 0.82) | 0.53 (0.21 to 0.86) | 0.004 |
| Schuetz *et al.*[11] | 0.84 (0.72 to 0.98) | 0.72 (0.54 to 0.96) | 0.52 (0.07 to 0.98) | 0.038 |
| Stone *et al.*[12] | 1.16 (0.97 to 1.40) | 1.35 (1.04 to 1.75) | 0.51 (0.07 to 0.94) | 0.025 |
| Vardeny *et al.*[13] | 1.06 (0.98 to 1.13) | 1.18 (0.88 to 1.59) | 0.33 (-0.31 to 0.96) | 0.038 |
| Lanz *et al.*[14] | 1.43 (1.06 to 1.92) | 2.96 (0.81 to 10.85) | 0.33 (-0.10 to 0.76) | 0.002 |
| Lomivorotov *et al.*[15] | 0.84 (0.66 to 1.06) | 0.52 (0.10 to 2.78) | 0.27 (-0.45 to 0.99) | 0.045 |
| Vermeersch *et al.*[16] | 0.84 (0.67 to 1.05) | 0.52 (0.13 to 2.06) | 0.27 (-0.35 to 0.88) | 0.019 |
| Frith *et al.*[17] | 1.28 (1.22 to 1.35) | 2.56 (1.35 to 4.83) | 0.26 (0.08 to 0.45) | <0.0001 |
| Wilson *et al.*[18] | 1.07 (0.86 to 1.33) | 1.36 (0.77 to 2.40) | 0.22 (-0.46 to 0.90) | 0.024 |
| Koch *et al.*[19] | 0.93 (0.84 to 1.02) | 0.65 (0.34 to 1.24) | 0.17 (-0.14 to 0.49) | <0.0001 |
| Willems *et al.*[20] | 1.21 (0.76 to 1.95) | 3.35 (0.70 to 16.11) | 0.16 (-0.22 to 0.55) | <0.0001 |
| Futier *et al.*[21] | 1.11 (0.91 to 1.36) | 2.00 (0.76 to 5.29) | 0.16 (-0.17 to 0.48) | <0.0001 |
| Onland *et al.*[22] | 0.95 (0.84 to 1.08) | 0.65 (0.42 to 0.99) | 0.11 (-0.17 to 0.39) | <0.0001 |
| Thiele *et al.*[23] | 1.03 (0.76 to 1.41) | 1.42 (0.46 to 4.40) | 0.09 (-0.77 to 0.94) | 0.036 |
| Karaye *et al.*[24] | 0.98 (0.80 to 1.20) | 0.39 (0.11 to 1.36) | 0.02 (-0.20 to 0.23) | <0.0001 |
| Lee *et al.*[25] | 1.00 (0.77 to 1.29) | 0.68 (0.36 to 1.30) | 0.01 (-0.66 to 0.67) | 0.003 |
| Araújo *et al.*[26] | 1.02 (0.76 to 1.36) | 0.41 (0.13 to 1.30) | -0.02 (-0.35 to 0.32) | <0.0001 |
| Zhang *et al.*[27] | 1.03 (0.60 to 1.78) | 0.57 (0.22 to 1.46) | -0.06 (-1.07 to 0.96) | 0.040 |
| De Luca *et al.*[28] | 0.95 (0.72 to 1.26) | 1.42 (0.76 to 2.66) | -0.14 (-1.08 to 0.80) | 0.017 |
| Tong *et al.*[29] | 0.64 (0.39 to 1.04) | 8.06 (1.01 to 64.15) | -0.22 (-0.57 to 0.14) | <0.0001 |
| Sekiziyivu *et al.*[30] | 0.91 (0.80 to 1.04) | 1.39 (0.56 to 3.44) | -0.27 (-1.16 to 0.62) | 0.005 |
| Johnston *et al.*[31] | 0.83 (0.72 to 0.97) | 1.33 (0.81 to 2.18) | -0.65 (-2.04 to 0.74) | 0.020 |

Abbreviations: RR<sub>c</sub>: relative risk for the composite outcome; RR<sub>d</sub>: relative risk for any-cause death; CI, confidence interval; BACO, bias attributable to composite outcomes; BACO index: $\ln(RR_c) / \ln(RR_d)$ .
<sup>a</sup>P value for the null hypothesis of the BACO index is equal to 1.

We observed that in 17 (73.9%) of these 23 studies, the BACO index value was between zero and <1; the remaining six had negative values, indicating a reversal of the association concerning a fatal event (mortality). None of the studies showed significant overestimation of the association attributable to the composite outcome (Table 3). More information on all 91 included articles can be found in the supplementary material.

### 3.4 Methodological quality of included studies

Of the 23 studies with significant or suggestive BACO, almost all had a low risk of bias (n=11) or some concerns (n=11). Only one article presented a high risk of bias, mainly due to the randomization process and outcome measurement (supplementary material).

## 4. Discussion

In our study, a quarter of the articles selected for review had a significant or suggestive BACO index. In all these cases, the BACO was significantly less than 1, indicating that the use of composite outcomes underestimated the association between the intervention and the prognosis, and in some cases even inverted the association.

The underestimation of the effect can be interpreted as a dilution of the association due to the inclusion of events affected differently by the intervention [6]. This phenomenon leads to a contradictory situation because composite outcomes are often used to obtain a higher number of events and, thus, greater statistical power. However, if the use of composite outcomes leads to a dilution of the effect, it paradoxically results in a reduction of the study’s power.

This explains situations such as the one observed in the study by Onland et al. [22], where hydrocortisone therapy was associated with a significant reduction in mortality among premature infants, while the effect on the composite outcome was not statistically significant (Table 3). Another example, recently recognized but beyond the scope of this review, is the study by Sesso et al. [32], where the use of a composite outcome underestimated the effect of cocoa extract supplementation on prognosis. This intervention did not reduce the incidence of the first cardiovascular event but was associated with a 27% reduction in mortality from that cause.

In another direction, underestimation can also lead to the failure to identify a harmful effect of the intervention. This occurred in the study by Stone et al. [12], where percutaneous coronary intervention (PCI) significantly increased mortality compared to coronary artery bypass grafting, while the association with the composite outcome was not statistically significant.

In other cases, the underestimation of the effect does not qualitatively change the conclusions but can lead to quantitatively significant differences, as seen in the study by Dangas et al. [9]. In this study, the effect of rivaroxaban was evaluated, finding that the excess risk for mortality was 67%. In contrast, the excess risk for the composite outcome was only 33%.

In six of the studies with a significant or suggestive BACO, the index value was negative, indicating an inversion of the association between the composite outcome and mortality. Beyond our review, we found an example of this trend in the study by Laurens et al. [33], which evaluated the effect of stopping prophylactic use of cotrimoxazole in adults with human immunodeficiency virus (HIV) infection. The authors compared standard prophylaxis using cotrimoxazole with its discontinuation, either alone or combined with chloroquine. The associations of the primary and secondary composite outcomes were in the opposite direction to mortality, with BACO indices of −0.48 (95% CI: −1.79 to 0.83; p = 0.03) and −0.69 (95% CI: −2.31 to 0.93; p = 0.04) for the primary and secondary composite outcomes, respectively [34]. This finding suggests that caution should be exercised when interpreting results, as they include effects in opposite directions. In such cases, it would be more appropriate to evaluate and interpret the components of the composite outcome individually.

There are some recommendations for constructing a composite outcome, such as considering the relevance of the outcomes for the patient, the frequency of the components of the composite outcome, and the similarity of the treatment effect for each of the events analyzed [3]. However, we believe it is challenging to construct a composite outcome that effectively represents the prognosis and allows for efficient evaluation of intervention effects. In this regard, the BACO index provides the possibility to test and guide interpretation, which can be predefined without affecting the study’s objectivity.

As limitations, our study is restricted to a relatively short time period and included primarily studies initiated before the pandemic. Therefore, we cannot infer that the prevalence and direction of BACO indices will be similar in other scenarios. However, a recent study focusing on clinical trials in COVID-19 patients estimated the BACO index in 28 effect estimates on composite outcomes [35]. In most studies, the composite outcome estimate was closer to the null value than that of mortality, and the BACO index was significantly less than 1 in five studies. Similar to the present study, there was no statistically significant overestimation of the effect associated with composite outcomes. Thus, we consider that in various scenarios, underestimation of the effect is likely the most prevalent bias resulting from the use of composite outcomes.

## 5. Conclusions

In a quarter of the selected studies, we observed suggestive results or statistically significant BACO. In most cases, the bias consisted of underestimating the effects, and in others, there was an inversion of the direction of the RR. These findings highlight the need to predefine guidelines for interpreting effects on composite endpoints based on objective criteria, such as the BACO index.

## CRediT authorship contribution statement

José Mário Nunes da Silva: Methodology, Supervision, Data curation, Formal analysis, Writing - original draft, Final approval of the version to be submitted. Juliana Ferreira Souza Conceição: Investigation, Formal analysis, Final approval of the version to be submitted. Paula C. Ramírez: Methodology, Investigation, Formal analysis, Writing - review & editing, Final approval of the version to be submitted. Christian Leonardo Diaz-León: Investigation, Formal analysis, Final approval of the version to be submitted. Fredi A. Diaz-Quijano: Conceptualization, Methodology, Project administration, Formal analysis, Writing - review & editing, Final approval of the version to be submitted.

## Supplementary data

Supplementary data for this article can be found in the appendix.

## Ethical statement

Local ethics review board approval was not required.

## Data Availability

All data produced in the present study are available upon reasonable request to the authors

## Supplementary Material

**Figure S1.**
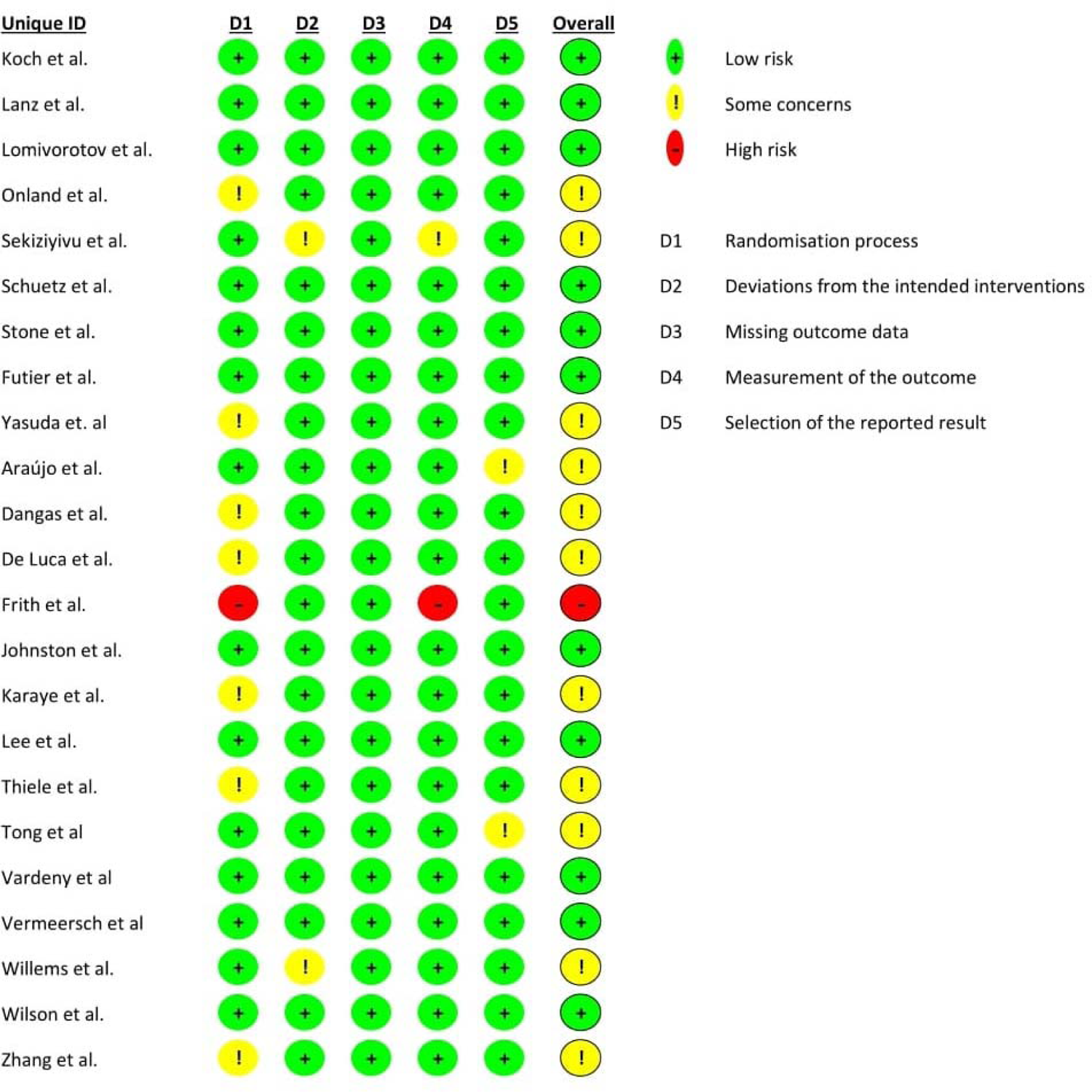
Assessment of bias risk in the 23 studies with significant or suggestive BACO.

